# Aspiration thrombectomy for patients with stroke and late onset to treatment: a subset analysis of the COMPLETE registry

**DOI:** 10.1101/2023.04.25.23288778

**Authors:** Ameer E. Hassan, Johanna T. Fifi, Osama O. Zaidat, COMPLETE Study Investigators

## Abstract

**Background:** The purpose of this study was to report the safety and performance of aspiration thrombectomy for patients with acute ischemic stroke (AIS) due to anterior circulation large vessel occlusion (LVO) and late onset to treatment.

**Methods:** This is a retrospective subset analysis of a global prospective multicenter registry (COMPLETE) that enrolled adults with AIS due to LVO and a pre-stroke modified Rankin Scale score (mRS) of 0 or 1 who were treated with aspiration thrombectomy with the Penumbra System. This subset analysis included all patients in the registry who had anterior circulation LVO, an Alberta Stroke Program Early CT Score of at least 6, and late onset to treatment (>6 hours from stroke onset to puncture).

**Results:** Of the 650 patients in the COMPLETE registry, 167 were included here. The rate of successful revascularization at the end of the procedure was 83.2% (139/167), the rate of good functional outcome (mRS 0-2) at 90 days was 55.4% (87/157), and the all-cause mortality rate at 90 days was 14.4% (24/167). No device-related serious adverse events (SAEs) occurred.

Procedure-related SAEs occurred in 9 patients (5.4%) within 24 hours and in 12 patients (7.2%) overall. No significant difference was detected between the outcomes of patients with an onset to puncture time of greater than 6 hours and less than or equal to 12 hours and the outcomes of patients with an onset to puncture time of greater than 12 hours.

**Conclusions:** For patients with AIS due to anterior circulation LVO and with late onset to treatment, aspiration thrombectomy with the Penumbra System appears to be safe and effective. The rates of good functional outcome and all-cause mortality from this study compared favorably with those rates from the medical management arms of the DAWN and DEFUSE-3 studies.

**Registration:** URL: https://www.clinicaltrials.gov; Unique identifier: NCT03464565.

## Introduction

In patients with acute ischemic stroke (AIS), a longer time from symptom onset to treatment, and thus a longer brain ischemia time, corresponds to a worse outcome.^1^ Additionally, with a longer time to treatment, the thrombus becomes more organized and potentially more compacted and adherent to the vessel wall.^2–4^ These thrombus characteristics could increase the difficulty, number of passes, and procedure time of mechanical thrombectomy,^5, 6^ thus increasing the chance of a complication. On the basis of 2 randomized controlled trials (the DAWN^7^ and DEFUSE-3^8^ studies) that compared mechanical thrombectomy plus medical management versus medical management alone in patients with AIS due to anterior circulation large vessel occlusion (LVO) and with a symptom onset to treatment or randomization time of greater than 6 hours, the current United States,^9^ European,^10^ and Society of Vascular and Interventional Neurology^11^ guidelines recommend treatment with mechanical thrombectomy in highly selected patients (per the DAWN and DEFUSE-3 studies) who present between 6 and 24 hours after symptom onset.

Patients in the DAWN and DEFUSE-3 studies were enrolled only if they had salvageable brain tissue as determined by using advanced imaging to measure infarcted brain tissue (CT perfusion or diffusion/perfusion MRI) and including only those patients with a small infarct volume and a mismatch between the infarct volume and the stroke severity (as measured by the National Institutes of Health Stroke Scale [NIHSS] score^7^ or the volume of the ischemic tissue^8^).^7, 8^ A recent registry study on endovascular thrombectomy patients who presented after 4.5 hours of symptom onset reported that patients who fulfilled the DAWN and DEFUSE-3 advanced imaging criteria had better outcomes and higher cost effectiveness than patients who did not.^12^

Two recent studies have been published on patients with AIS due to anterior circulation LVO and late onset to treatment who were treated with mechanical thrombectomy, with aspiration thrombectomy used in some but not all patients.^13, 14^ Neither study used the selection criteria in the DAWN and DEFUSE-3 studies, and neither study detected a significant difference in outcomes between patients with early versus late onset to treatment.^13, 14^ No previous studies are available that report the results of aspiration thrombectomy in all patients for the treatment of AIS with late onset to treatment. The purpose of this study was to report the safety and performance of aspiration thrombectomy with the Penumbra System (Penumbra, Inc., Alameda) for patients with AIS due to anterior circulation LVO and with late onset to treatment, in a real-world setting.

## Methods

This study is a retrospective subset analysis of a global prospective multicenter observational registry that included patients who presented with either anterior or posterior LVO and were eligible for aspiration thrombectomy using the Penumbra System including the Penumbra 3D Revascularization Device (Penumbra, Inc.). This registry, COMPLETE (International Acute Ischemic Stroke Registry with the Penumbra System Aspiration Including the 3D Revascularization Device), was registered with ClinicalTrials.gov (NCT03464565). The study and the informed consent process were approved by the Institutional Review Board/Ethics Committee (IRB/EC) for each participating center. The enrollment period was July 2018 through October 2019, and 90-day follow-up was completed January 2020. This subset analysis included all patients in the registry who had anterior circulation LVO, an Alberta Stroke Program Early CT Score (ASPECTS) of at least 6, and late onset to treatment (>6 hours from stroke onset to puncture).

The study protocol for the COMPLETE registry is described in detail elsewhere.^15^ Patients were included if they were at least 18 years old, had a pre-stroke modified Rankin Scale score (mRS) of 0 or 1, experienced AIS secondary to intracranial LVO and eligible for mechanical thrombectomy using the Penumbra System, had planned frontline treatment with the Penumbra System, and had signed informed consent per the center’s IRB/EC. Enrolled patients were treated with aspiration thrombectomy either alone or in combination with the Penumbra 3D Revascularization Device. Primary endpoints were good functional outcome (mRS 0-2) at 90 days, all-cause mortality at 90 days, and successful revascularization (modified thrombolysis in cerebral infarction score [mTICI] 2b-3 achieved) at the end of the procedure. Secondary endpoints were successful revascularization after the first pass, device-related and procedure-related serious adverse events (SAEs), embolization in new or uninvolved territory as seen on the final angiogram, symptomatic intracranial hemorrhage (sICH) within 24 hours, vessel perforation, and vessel dissection. Imaging findings were evaluated by a core lab, and clinical events related to safety endpoints were adjudicated by independent medical reviewers.

Data analyses were performed by using SAS (version 9.4, SAS Institute). Descriptive statistics were calculated for all patients and for the subgroups of patients with an onset to puncture time of greater than 6 hours and less than or equal to 12 hours (6-12h subgroup) and patients with an onset to puncture time of greater than 12 hours (>12h subgroup). The 2 subgroups were also compared by using the 2-tailed t-test, Mann-Whitney test, or Fisher exact test as appropriate to calculate *P*-values.

## Results

Of the 650 patients in the COMPLETE registry, 167 patients with anterior LVO, an ASPECTS of at least 6, and a late onset to treatment time (>6 hours) were included in this subset analysis (Table 1). Median patient age was 70 years (IQR 61-78), and 94 patients (56.3%) were female. Race and ethnicity data were collected only for the 119 patients (71.3%) in the United States; most patients in the United States were White (94/119, 79.0%) or Black or African American (18/119, 15.1%). The most common medical history characteristics were hypertension (120, 71.9%), cardiovascular/vascular disease (80, 47.9%), and hyperlipidemia (73, 43.7%). Stroke onset was unwitnessed in 85 patients (51.2%), present upon wakeup in 43 patients (25.9%), and witnessed in 38 patients (22.9%). One hundred and eight (64.7%) patients were transferred from another hospital. The median time from stroke onset to hospital admission was 8.6 hours (IQR 6.1-13.1). Intravenous tissue plasminogen activator (IV tPA) was administered before the procedure in 36 patients (21.6%); most of those patients (30, 83.3%) were transferred from another hospital.

**Table 1.**
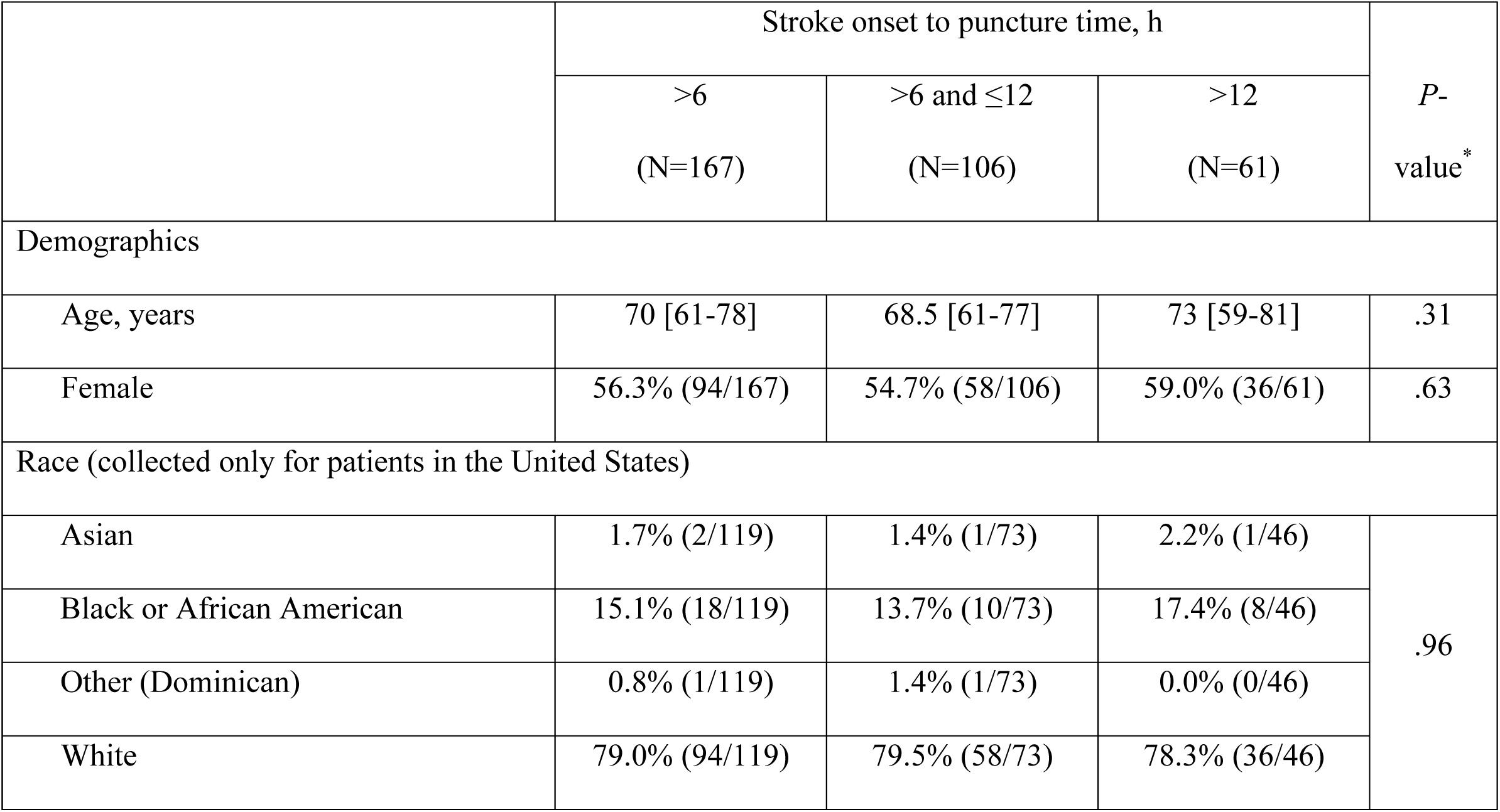
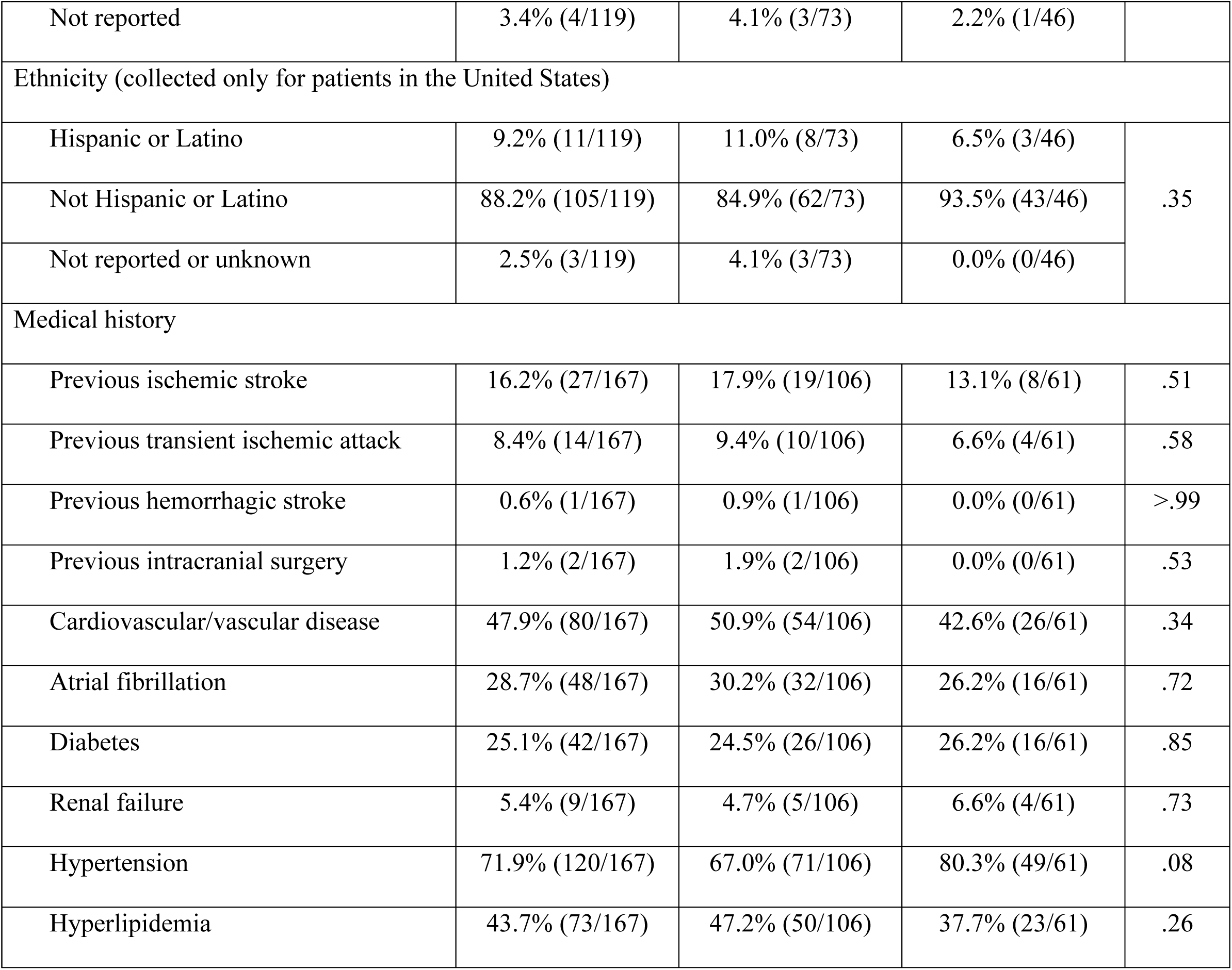
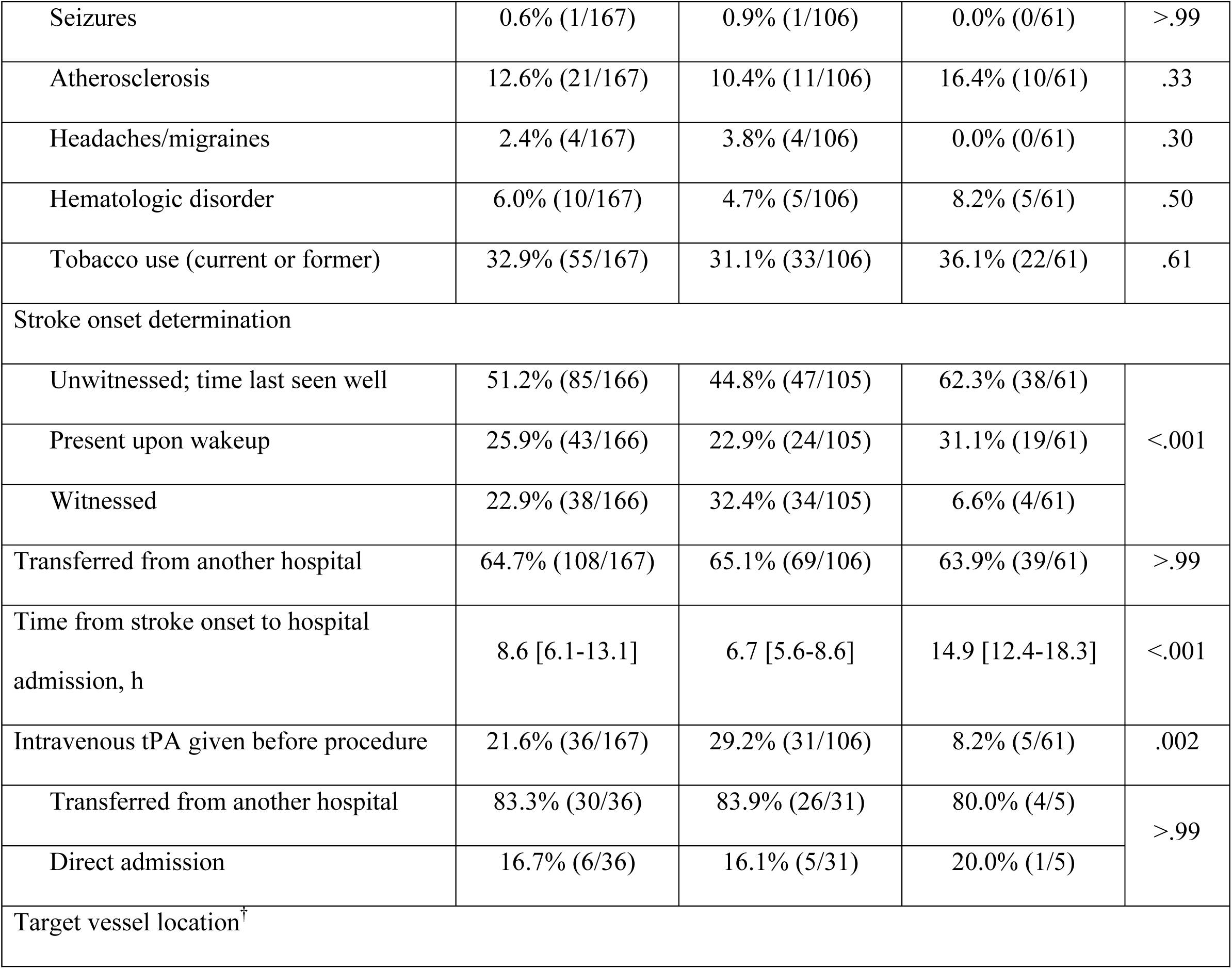
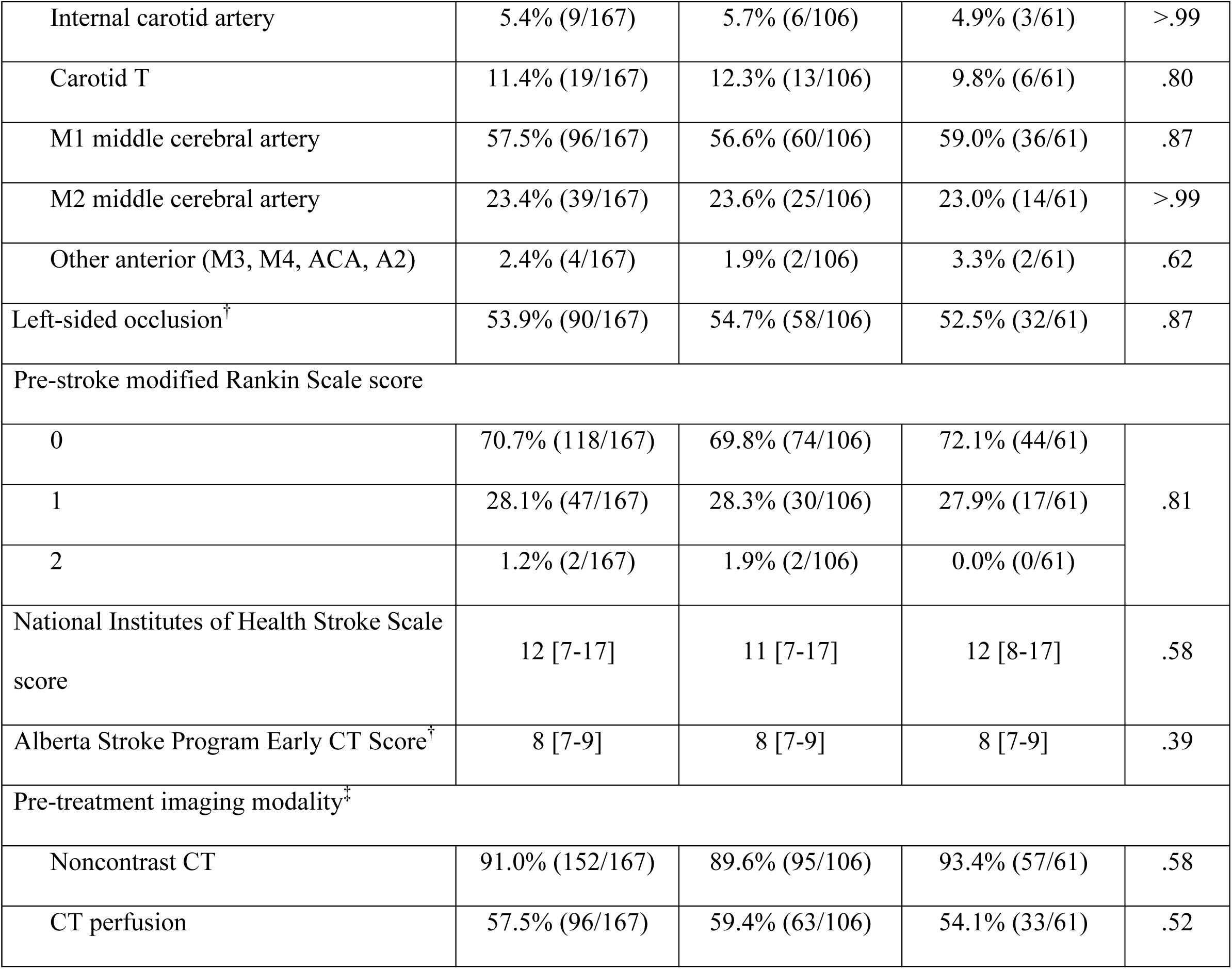
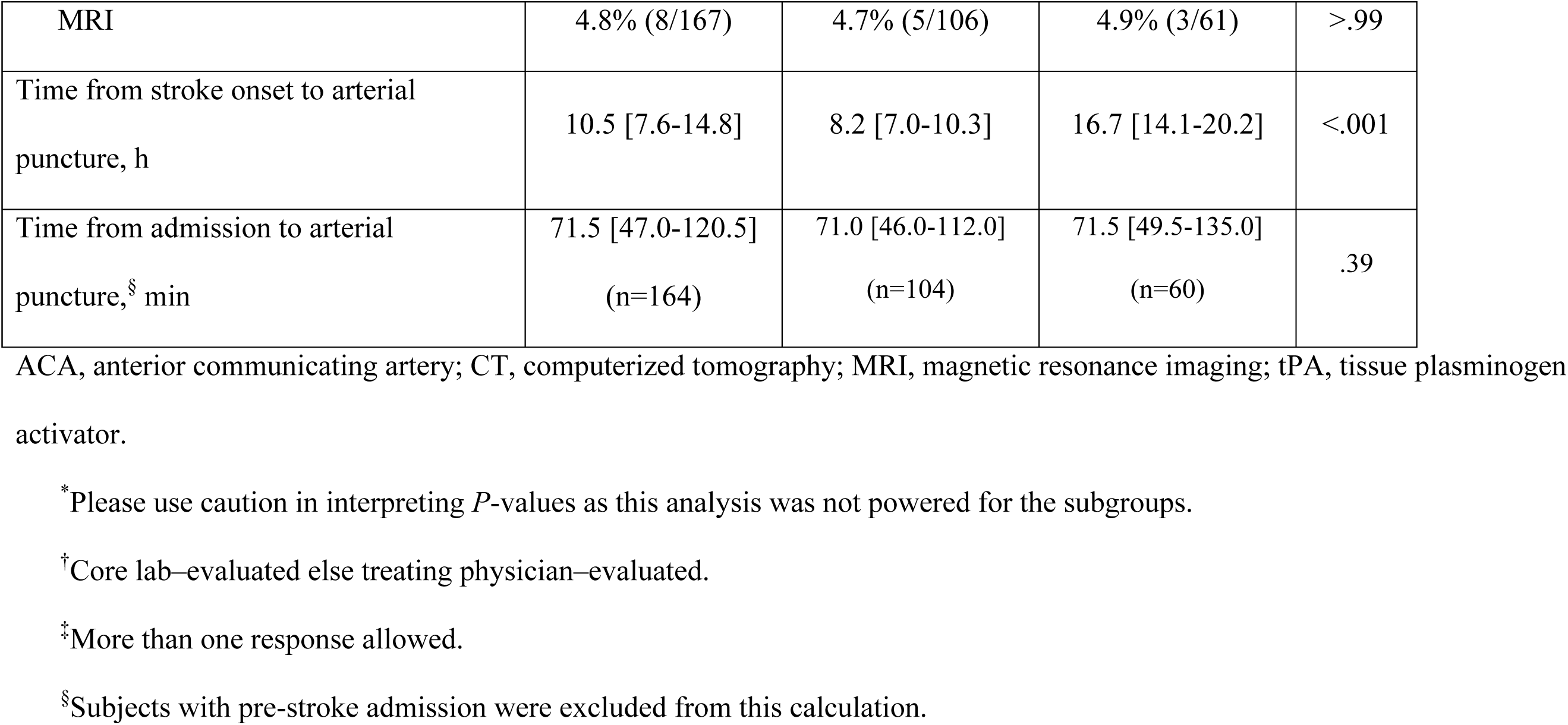
Baseline characteristics and presentation for patients treated with aspiration thrombectomy for acute ischemic stroke due to anterior circulation large vessel occlusion and with late onset to treatment (>6 hours from stroke onset to puncture). Continuous variables are reported as median [IQR] and categorical variables are reported as percentage (n/N).

The most common target vessel location was the M1 middle cerebral artery (96, 57.5%; Table 1). The occlusion was on the left side in 90 patients (53.9%). Pre-stroke mRS was 0 in 118 patients (70.7%), 1 in 47 patients (28.1%), and 2 in 2 patients (1.2%). The median pre-procedure NIHSS score was 12 (IQR 7-17). The median pre-procedure ASPECTS was 8 (IQR 7-9). Patients were assessed by using noncontrast CT (152/167, 91.0%), CT perfusion (96/167, 57.5%) and MRI (8/167, 4.8%). The median time from stroke onset to arterial puncture was 10.5 hours (IQR 7.6-14.8), and the median time from hospital admission to arterial puncture was 71.5 minutes (IQR 47.0-120.5, N=164).

The frontline procedural technique was aspiration alone in 100 patients (59.9%), aspiration plus the 3D Revascularization Device in 64 patients (38.3%), and other in 3 patients (1.8%, Table 2). A single pass was performed in 64 patients (38.3%). The median time from stroke onset to mTICI 2b-3 or to the final angiogram was 11.1 hours (IQR 8.4-15.7). The median time from arterial puncture to mTICI 2b-3 or to the final angiogram was 28 minutes (IQR 17-47).

**Table 2.** Procedural information for patients treated with aspiration thrombectomy for acute ischemic stroke due to anterior circulation large vessel occlusion and with late onset to treatment (>6 hours from stroke onset to puncture). Continuous variables are reported as median [IQR] and categorical variables are reported as percentage (n/N).

|  | Stroke onset to puncture time, h |  |  | *P*-value<sup>*</sup> |
| --- | --- | --- | --- | --- |
|  | >6<br>(N=167) | >6 and ≤12<br>(N=106) | >12<br>(N=61) |  |
| Frontline procedural technique |  |  |  |  |
| Aspiration alone | 59.9% (100/167) | 62.3% (66/106) | 55.7% (34/61) | .74 |
| Aspiration plus 3D | 38.3% (64/167) | 35.8% (38/106) | 42.6% (26/61) |  |
| Other | 1.8% (3/167) | 1.9% (2/106) | 1.6% (1/61) |  |
| Number of passes |  |  |  |  |
| 1 | 38.3% (64/167) | 46.2% (43/93) | 38.2% (21/55) | .66 |
| 2 | 21.6% (36/167) | 23.7% (22/93) | 25.5% (14/55) |  |
| 3 | 20.4% (34/167) | 22.6% (21/93) | 23.6% (13/55) |  |
| 4 or more | 19.8% (33/167) | 7.5% (7/93) | 12.7% (7/55) |  |
|  | >6<br>(N=167) | >6 and ≤12<br>(N=106) | >12<br>(N=61) |  |
| Time from stroke onset to mTICI 2b-3 or final angiogram, h<sup>†</sup> | 11.1 [8.4-15.7] | 9.0 [7.6-10.8] | 17.1 [14.9-20.5] | <.001 |
| Time from arterial puncture to mTICI 2b-3 or final angiogram, min<sup>†</sup> | 28 [17-47] | 31 [17-47] | 25 [17-42] | .65 |

3D, 3D Revascularization Device; mTICI, modified treatment in cerebral infarction score.
<sup>\*</sup>Please use caution in interpreting *P*-values as this analysis was not powered for the subgroups.
<sup>†</sup>Core lab–evaluated.

The rate of successful revascularization at the end of the procedure was 83.2% (139/167), the rate of good functional outcome at 90 days was 55.4% (87/157), and the all-cause mortality rate at 90 days was 14.4% (24/167, Table 3). The rate of successful revascularization after the first pass was 49.1% (81/165). No device-related SAEs occurred. Procedure-related SAEs occurred in 9 patients (5.4%) within 24 hours and in 12 patients (7.2%) overall. Embolization in new or uninvolved territory at the end of the procedure occurred in 4 patients (2.4%) and sICH within 24 hours occurred in 7 patients (4.2%). Vessel perforation occurred in 0 patients and vessel dissection occurred in 1 patient (0.6%).

**Table 3.**
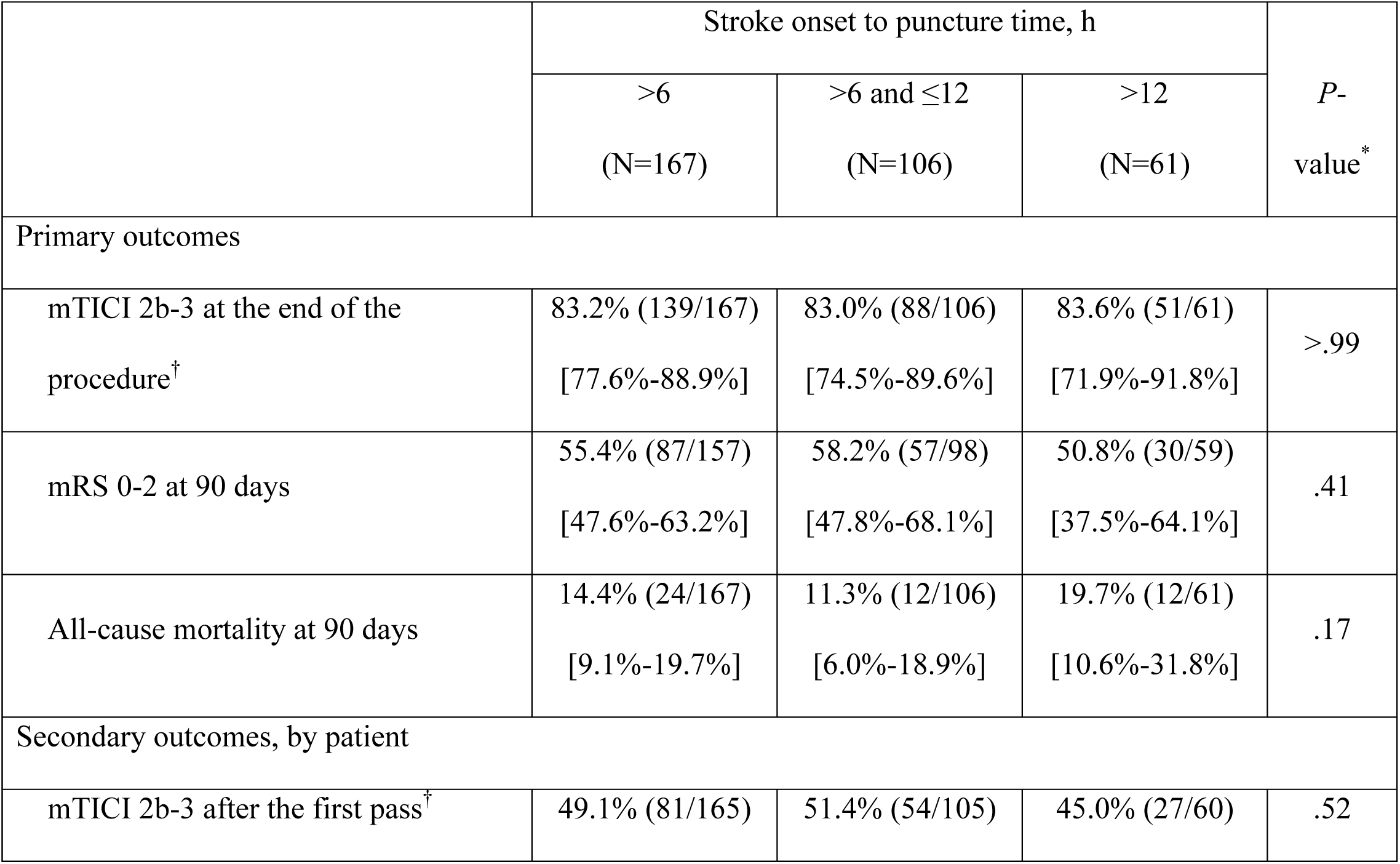
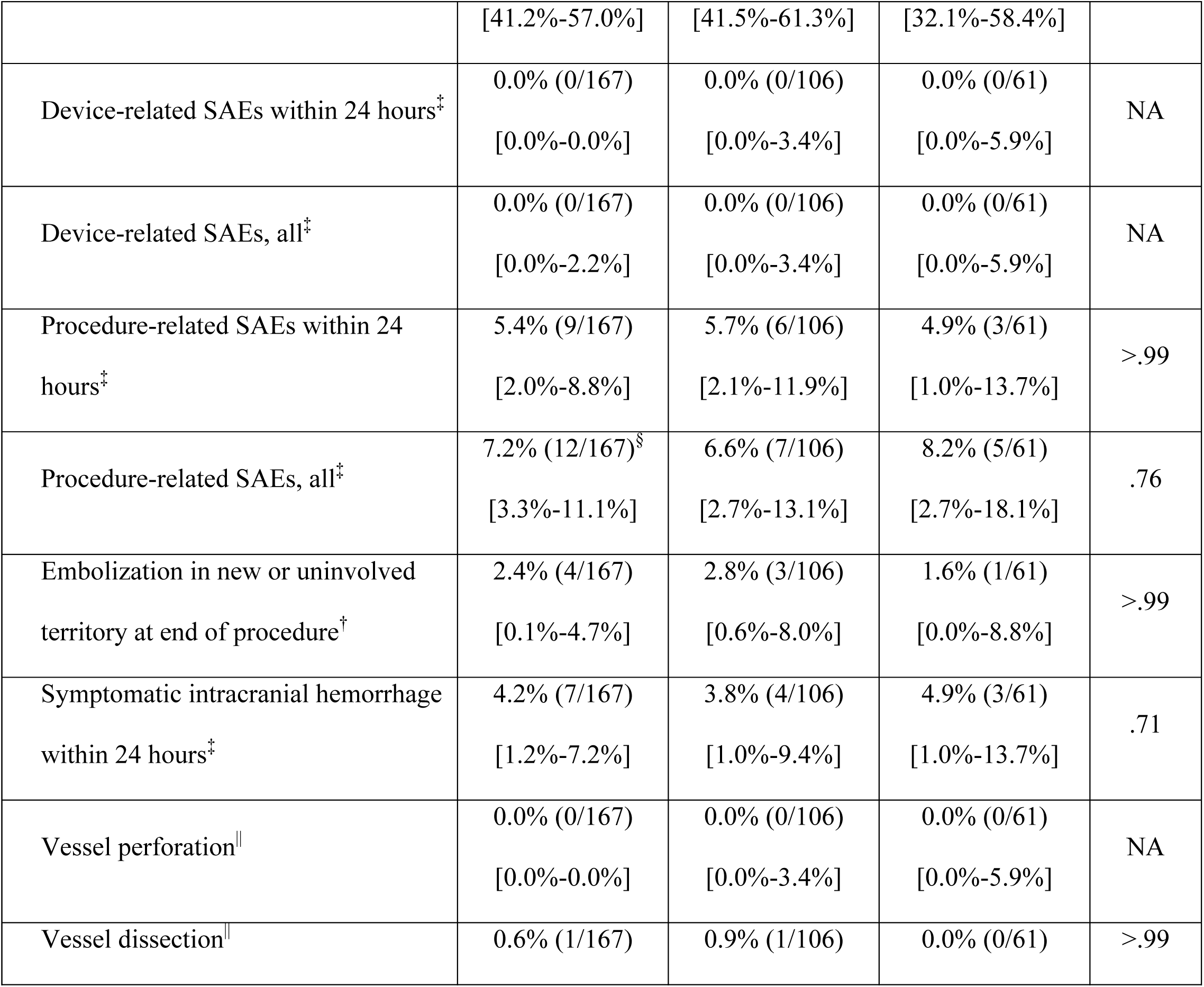
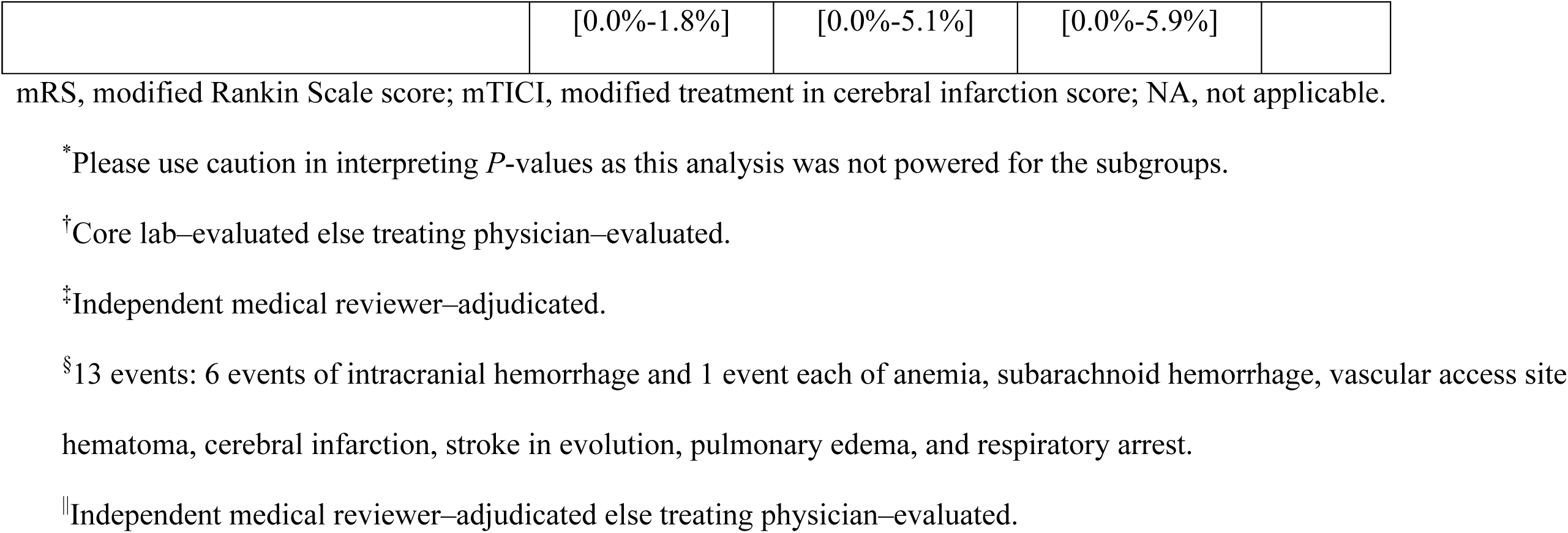
Outcomes for patients treated with aspiration thrombectomy for acute ischemic stroke due to anterior circulation large vessel occlusion and with late onset to treatment (>6 hours from stroke onset to puncture). Variables are reported as percentage (n/N) [95% CI].

The median time from stroke onset to hospital admission was 6.7 hours for the 6-12h subgroup and 14.9 hours for the >12h subgroup (*P* <.001), and the median time from stroke onset to arterial puncture was 8.2 hours for the 6-12h subgroup and 16.7 hours for the >12h subgroup (*P* <.001; Table 1). A statistically significant difference was observed between the subgroups for the distribution of stroke onset types (*P* <.001); higher proportions of patients in the >12h subgroup had a stroke onset that was unwitnessed or that was present upon wakeup. Otherwise, no significant difference was detected between the 2 subgroups for any baseline characteristics except for IV tPA given before procedure (6-12h subgroup, 29.2%; >12h subgroup, 8.2%; *P* =.002; Table 1). No significant difference was detected between the 2 subgroups for any procedural characteristics except for time from stroke onset to mTICI 2b-3 or final angiogram (6-12h subgroup, 9.0 hours; >12h subgroup, 17.1 hours; *P* <.001; Table 2). For all outcomes, no significant difference was detected between the 2 subgroups (Table 3).

## Discussion

Patients with AIS due to anterior circulation LVO and with late onset to treatment can be effectively treated with aspiration thrombectomy. Late onset to treatment patients selected per the DAWN and DEFUSE-3 advanced imaging criteria are more likely to benefit from mechanical thrombectomy,^12^ and current guidelines on treating late onset to treatment stroke patients with mechanical thrombectomy^9–11^ are based on the results of DAWN and DEFUSE-3 studies. In real-world situations, however, use of advanced imaging to determine whether patients with late onset to treatment have salvageable brain tissue is challenging at some locations, as advanced imaging is less available, more complex, and more costly than simpler imaging modalities; can delay treatment; can increase exposure to contrast medium and radiation; and can exclude patients who might benefit from mechanical thrombectomy despite not meeting the stringent inclusion criteria from the DAWN and DEFUSE-3 studies.^16, 17^ Other stroke patients with late onset to treatment can benefit from mechanical thrombectomy, including those patients with signs of less severe stroke, especially with a higher ASPECTS^13, 14, 16, 18^ or good collateral status.^18–20^ In this study, patients with late onset to treatment, anterior circulation LVO, and an ASPECTS of at least 6 had a high rate of successful revascularization, a high rate of good functional outcome at 90 days, and a low mortality rate at 90 days. Additionally, no significant difference was detected between the outcomes of patients with an onset to puncture time of greater than 6 hours and less than or equal to 12 hours and the outcomes of patients with an onset to puncture time of greater than 12 hours. This study presents real-world experience of treating patients with AIS due to anterior circulation LVO and with late onset to treatment.

The rates of good functional outcome at 90 days and of all-cause mortality at 90 days from this study compared favorably with those rates from the medical management arms of the DAWN and DEFUSE-3 studies (Table 4).^7, 8^ The rate of good functional outcome at 90 days was considerably higher in the current study (55.4%) than in the medical management arms of the DAWN (13.1%) and DEFUSE-3 (16.7%) studies, and the all-cause mortality rate at 90 days was lower in the current study (14.4%) than in the medical management arms of the DAWN (18.2%) and DEFUSE (25.6%) studies. Additionally, the rate of good functional outcome at 90 days was higher in the current study (55.4%) than in the stent retriever thrombectomy arm of the DAWN study (48.6%) and in the mechanical thrombectomy arm of the DEFUSE-3 study (44.6%). The all-cause mortality rate at 90 days in the current study (14.4%) was comparable to the rates in the stent retriever thrombectomy arm of the DAWN study (18.7%) and the mechanical thrombectomy arm of the DEFUSE-3 study (14.1%), and the sICH rate in the current study (4.2%) was comparable to the rates in the stent retriever thrombectomy arm of the DAWN study (5.6%) and the mechanical thrombectomy arm of the DEFUSE-3 study (6.5%).

**Table 4.** Comparison of the current study (from the COMPLETE registry) with the DAWN and DEFUSE-3 studies,^7, 8^ for patients with acute ischemic stroke due to anterior circulation large vessel occlusion and with late onset to treatment who were treated with mechanical thrombectomy plus medical management or with medical management. Continuous variables are reported as mean (SD) or as median [IQR] and categorical variables are reported as percentage (n/N).

| Study or registry name | Treatment | Endpoints |  |  |
| --- | --- | --- | --- | --- |
|  |  | mRS 0-2 at 90 days | All-cause mortality at 90 days | sICH |
| COMPLETE | Aspiration thrombectomy ± 3D | 55.4%<br>(87/157) | 14.4%<br>(24/167) | 4.2%<br>(7/167) |
| DAWN<sup>7</sup> | Stent retriever thrombectomy + medical management | 48.6%<br>(52/107) | 18.7%<br>(20/107) | 5.6%<br>(6/107) |
|  | Medical management | 13.1%<br>(13/99) | 18.2%<br>(18/99) | 3.0%<br>(3/99) |
|  | *P*-value | >.99* | NSD | NSD |
| DEFUSE-3<sup>8</sup> | Mechanical thrombectomy + medical management | 44.6%<br>(41/92) | 14.1%<br>(13/92) | 6.5%<br>(6/92) |
|  | Medical management | 16.7%<br>(15/90) | 25.6%<br>(23/90) | 4.4%<br>(4/90) |

| Study or<br>registry name | Treatment | Endpoints |  |  |
| --- | --- | --- | --- | --- |
|  |  | mRS 0-2<br>at 90 days | All-cause<br>mortality at<br>90 days | sICH |
|  | *P*-value | <.001 | .05 | .75 |

3D, 3D Revascularization Device; mRS, modified Rankin Scale score;
NSD, no significant difference; sICH, symptomatic intracranial
hemorrhage.
\*Posterior probability of superiority.

The outcomes from the current study were in range of the outcomes reported by other studies of late onset to treatment in patients with AIS due to anterior circulation LVO (Supplemental Table S1).^7, 8, 13, 14, 16–18, 21–24^ The rate of good functional outcome was considerably higher for all the mechanical thrombectomy study arms or studies (including the current study) than for the all the medical management arms among the studies. Heterogeneity among these studies in regard to study design, baseline NIHSS, imaging and subsequent patient selection, percentage of patients with M2 middle cerebral artery occlusion, IV tPA given before the procedure, and treatment modality limits direct comparisons between them. In particular, the median NIHSS was appreciably lower and the rate of IV tPA given before the procedure was appreciably higher in the current study than in the DAWN and DEFUSE-3 studies.^7, 8^

A limitation of this study is that it was a post hoc analysis of prospectively collected data. This limitation is reflective of the real-world situation in which the data for this study were collected. The statistical analysis was not powered for the 2 subgroups. The proportions of patients who had a stroke onset that was unwitnessed or that was present upon wakeup were higher for the >12h subgroup than in the 6-12h subgroup, but those stroke onset types would most likely result in a longer time to treatment. The rate of IV tPA given before the procedure was statistically significant between the 2 subgroups and higher in the 6-12h subgroup; current guidelines recommend IV tPA only in selected patients within 3 and 4.5 hours of onset^9^ or within 6 hours of onset, so IV tPA would be more likely to have been administered in patients with a shorter onset to puncture time.^10^ Sites might have measured infarcted brain tissue by using advanced imaging (per DAWN or DEFUSE-3 guidelines) to determine patient eligibility as part of the sites’ standard of care; however, neither DAWN nor DEFUSE-3 criteria were inclusion criteria for this study. The study population was limited to patients with anterior circulation LVO and an ASPECTS of at least 6; however, other studies on AIS with late onset to treatment have similarly limited their study population to patients with anterior circulation LVO, and most of those studies (excepting 1 study on patients with an ASPECTS of 5 or less^24^ and the DAWN study, which did not report ASPECTS^7^) reported a median ASPECTS that was identical to or at most 1 unit higher than that reported in the current study (Supplemental Table S1).^8, 13, 14, 16–18, 21–23^

In conclusion, for patients with acute ischemic stroke due to anterior circulation large vessel occlusion, and with late onset to treatment, aspiration thrombectomy with the Penumbra System was safe and effective.

## Data Availability

No statement.

## Non-standard Abbreviations and Acronyms

ACA: Anterior communicating artery
AIS: Acute ischemic stroke
ASPECTS: Alberta Stroke Program Early CT Score
IRB/EC: Institutional Review Board/Ethics Committee
LVO: Large vessel occlusion
MCA: Middle cerebral artery
mRS: Modified Rankin Scale score
mTICI: Modified thrombolysis in cerebral infarction score
NIHSS: National Institutes of Health Stroke Scale
sICH: Symptomatic intracranial hemorrhage

## Acknowledgements

The authors acknowledge the contributions of all research coordinators and investigators that contributed to the COMPLETE (International Acute Ischemic Stroke Registry With the Penumbra System Aspiration Including the 3D Revascularization Device) Registry’s data collection and entry process. The authors additionally acknowledge Penumbra employees Hee Jung Lee, MS, and Nam Nguyen, MS, for statistical assistance, and Anneliese D. Heiner, PhD, for writing assistance.

## Supplemental Material

Table S1

## Tables

**Supplemental Table S1.**
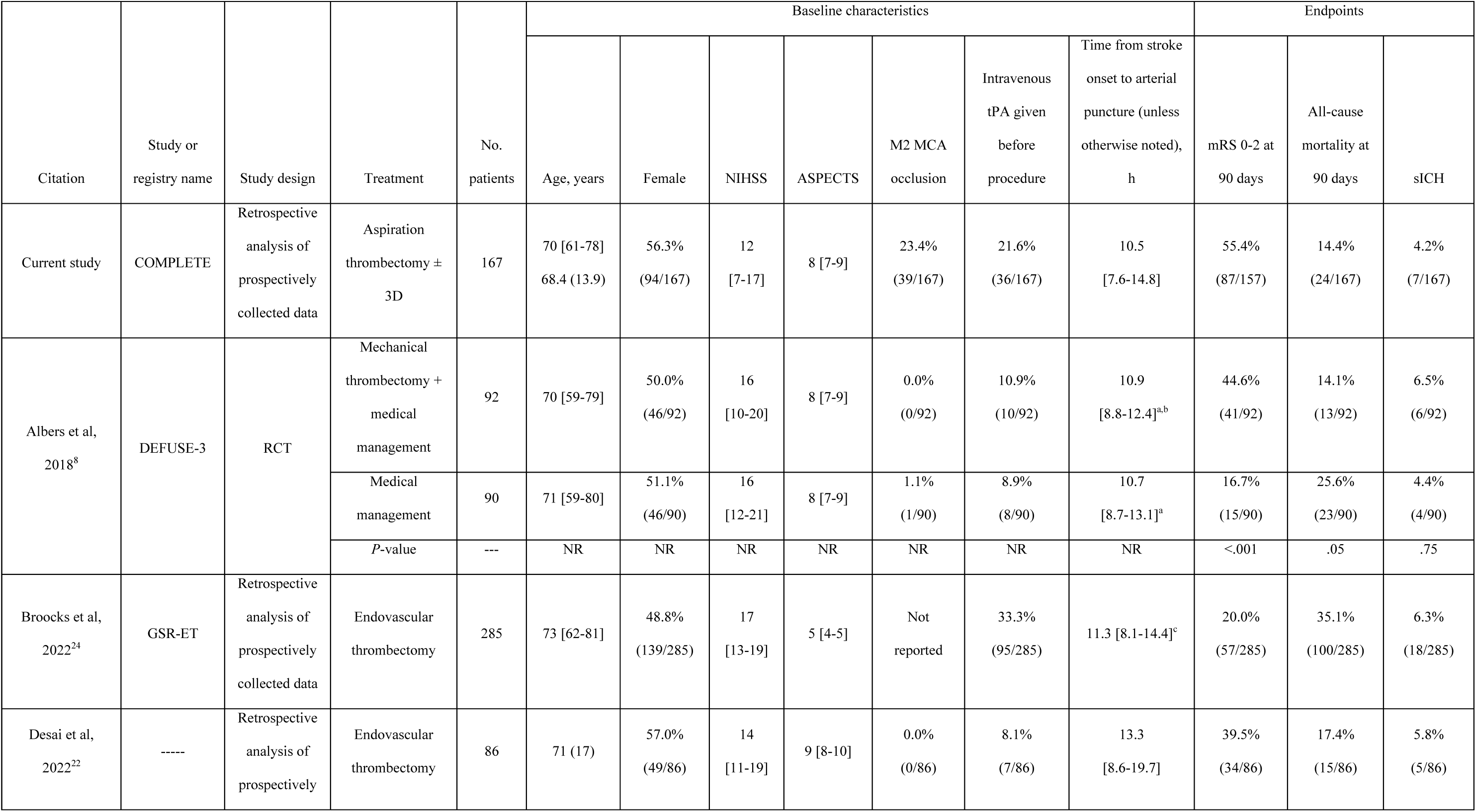
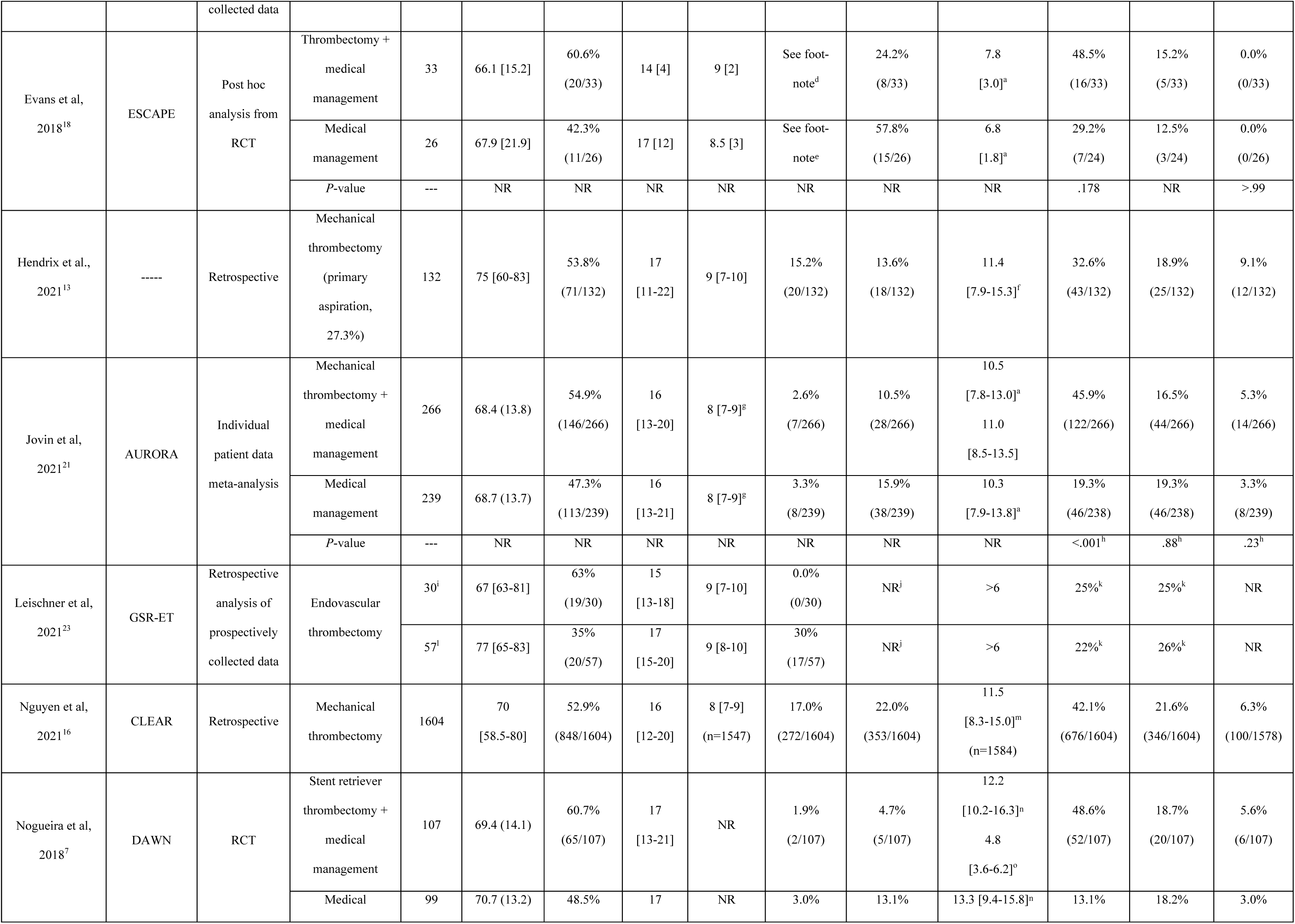
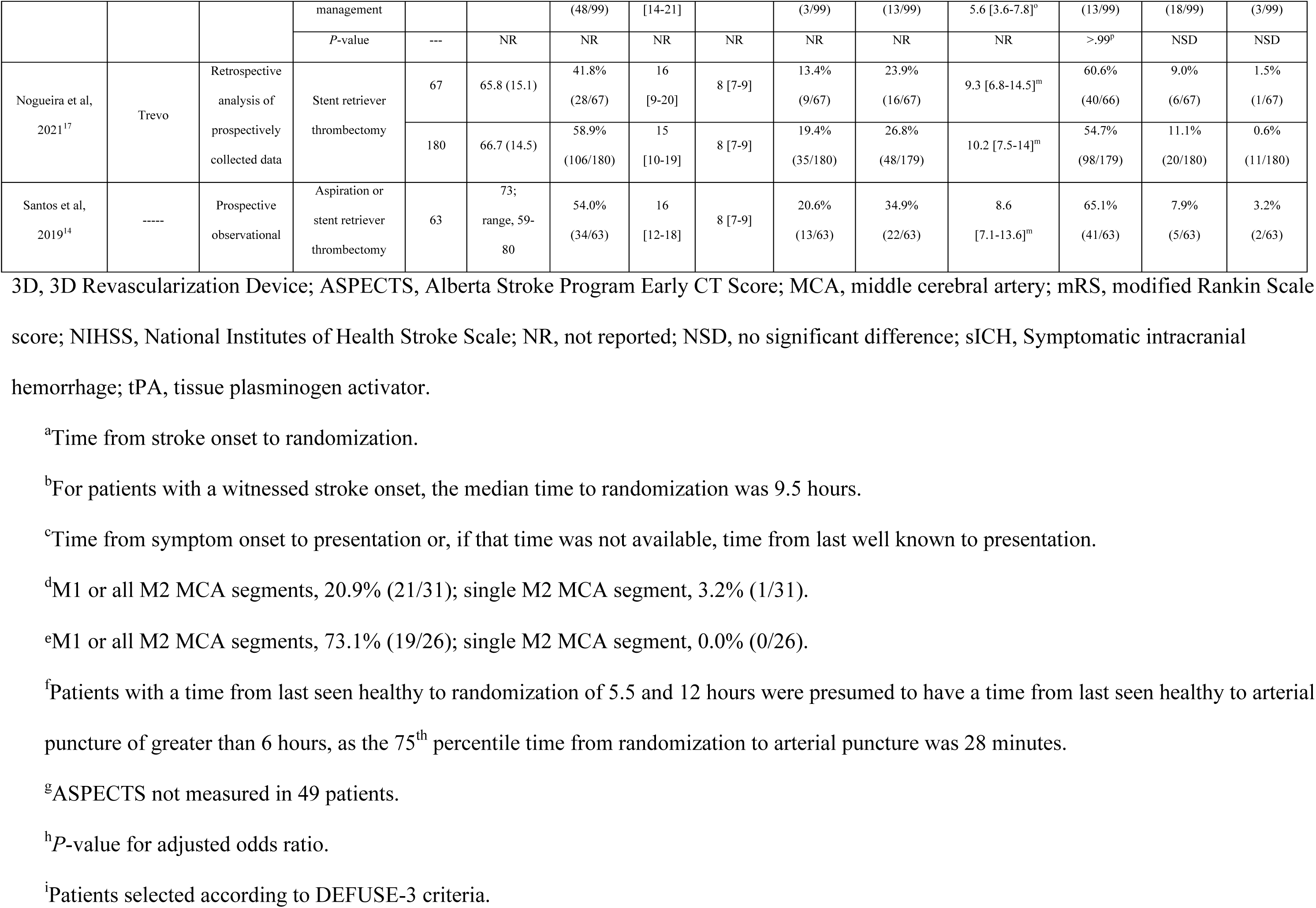
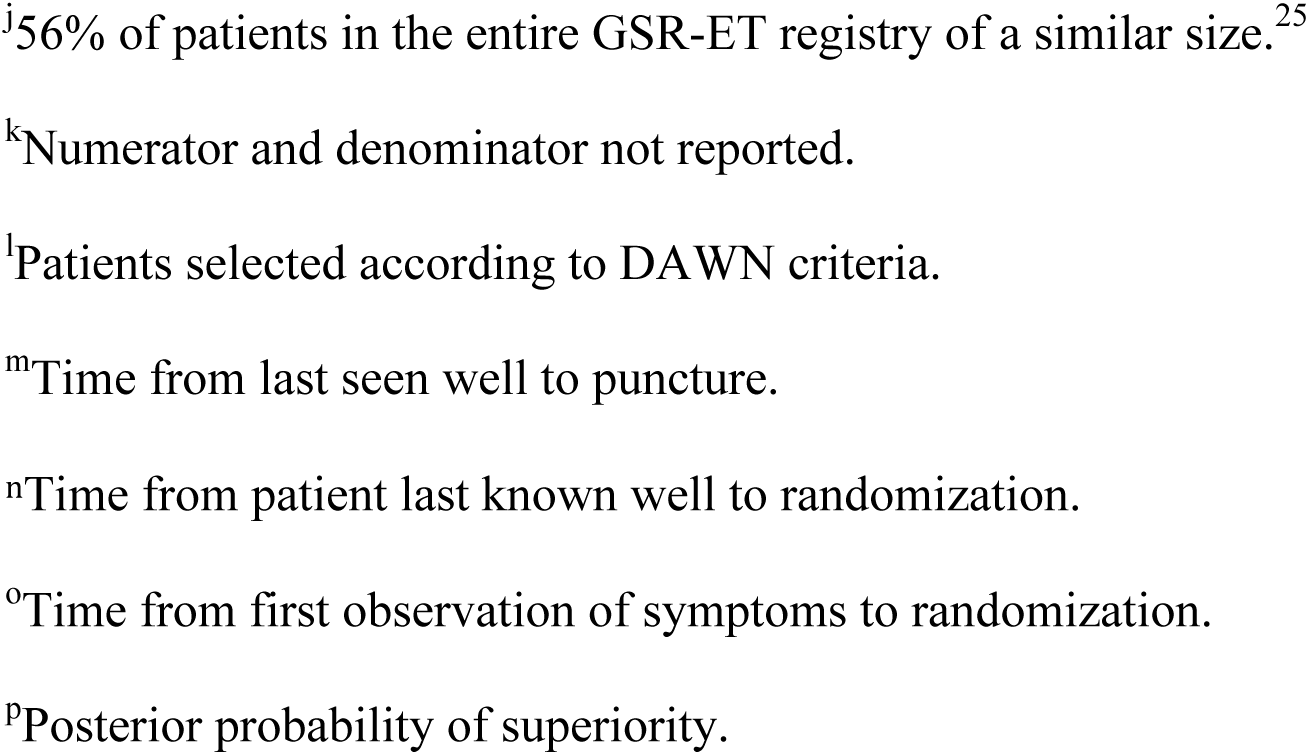
Comparison of patients with acute ischemic stroke due to anterior circulation large vessel occlusion and with late onset to treatment who were treated with mechanical thrombectomy plus medical management or with medical management. Continuous variables are reported as mean (SD) or as median [IQR] and categorical variables are reported as percentage (n/N).

## References

1. Almekhlafi MA, Goyal M, Dippel DWJ, Majoie CBLM, Campbell BCV, Muir KW, et al. Healthy life-year costs of treatment speed from arrival to endovascular thrombectomy in patients with ischemic stroke: A meta-analysis of individual patient data from 7 randomized clinical trials. JAMA Neurology. 2021;78:709–717

2. Donnerstag F, Götz F, Dadak M, Raab P, Iglesias EC, Werlein C, et al. Interventional stroke treatment – is it also safe for arteries? Looking at thrombectomy wall damage through clot histology. Interventional Neuroradiology. 2021;27:404–410

3. Yoo AJ, Andersson T. Thrombectomy in acute ischemic stroke: Challenges to procedural success. Journal of Stroke. 2017;19:121–130

4. Jolugbo P, Ariëns RAS. Thrombus composition and efficacy of thrombolysis and thrombectomy in acute ischemic stroke. Stroke. 2021:1131–1142

5. Shimizu H, Hatakeyama K, Saito K, Shobatake R, Takahashi N, Deguchi J, et al. Age and composition of the thrombus retrieved by mechanical thrombectomy from patients with acute ischemic stroke are associated with revascularization and clinical outcomes. Thrombosis Research. 2022;219:60–69

6. Kitano T, Hori Y, Okazaki S, Shimada Y, Iwamoto T, Kanki H, et al. An older thrombus delays reperfusion after mechanical thrombectomy for ischemic stroke. Thrombosis and Haemostasis. 2022;122:415–426

7. Nogueira RG, Jadhav AP, Haussen DC, Bonafe A, Budzik RF, Bhuva P, et al. Thrombectomy 6 to 24 hours after stroke with a mismatch between deficit and infarct. New England Journal of Medicine. 2018;378:11–21

8. Albers GW, Marks MP, Kemp S, Christensen S, Tsai JP, Ortega-Gutierrez S, et al. Thrombectomy for stroke at 6 to 16 hours with selection by perfusion imaging. New England Journal of Medicine. 2018;378:708–718

9. Powers WJ, Rabinstein AA, Ackerson T, Adeoye OM, Bambakidis NC, Becker K, et al. 2018 guidelines for the early management of patients with acute ischemic stroke: A guideline for healthcare professionals from the american heart association/american stroke association. Stroke. 2018;49:e46–e110

10. Turc G, Bhogal P, Fischer U, Khatri P, Lobotesis K, Mazighi M, et al. European stroke organisation (eso) - european society for minimally invasive neurological therapy (esmint) guidelines on mechanical thrombectomy in acute ischemic stroke. Journal of NeuroInterventional Surgery. 2019

11. Nguyen TN, Castonguay AC, Siegler JE, Nagel S, Lansberg MG, de Havenon A, et al. Mechanical thrombectomy in the late presentation of anterior circulation large vessel occlusion stroke: A guideline from the society of vascular and interventional neurology guidelines and practice standards committee. Stroke: Vascular and Interventional Neurology. 2022:e000512

12. Gao L, Bivard A, Parsons M, Spratt NJ, Levi C, Butcher K, et al. Real-world cost-effectiveness of late time window thrombectomy for patients with ischemic stroke. Frontiers in Neurology. 2021;12

13. Hendrix P, Chaudhary D, Avula V, Abedi V, Zand R, Noto A, et al. Outcomes of mechanical thrombectomy in the early (<6-hour) and extended (≥6-hour) time window based solely on noncontrast ct and ct angiography: A propensity score-matched cohort study. American Journal of Neuroradiology. 2021;42

14. Santos T, Carvalho A, Cunha AA, Rodrigues M, Gregório T, Paredes L, et al. Ncct and cta-based imaging protocol for endovascular treatment selection in late presenting or wake-up strokes. Journal of NeuroInterventional Surgery. 2019;11:190–195

15. Zaidat OO, Fifi JT, Nanda A, Atchie B, Woodward K, Doerfler A, et al. Endovascular treatment of acute ischemic stroke with the penumbra system in routine practice: Complete registry results. Stroke. 2021:STROKEAHA121034268

16. Nguyen TN, Abdalkader M, Nagel S, Qureshi MM, Ribo M, Caparros F, et al. Noncontrast computed tomography vs computed tomography perfusion or magnetic resonance imaging selection in late presentation of stroke with large-vessel occlusion. JAMA Neurology. 2022;79:22–31

17. Nogueira RG, Haussen DC, Liebeskind D, Jovin TG, Gupta R, Jadhav A, et al. Stroke imaging selection modality and endovascular therapy outcomes in the early and extended time windows. Stroke. 2021:491–497

18. Evans JW, Graham BR, Pordeli P, Al-Ajlan FS, Willinsky R, Montanera WJ, et al. Time for a time window extension: Insights from late presenters in the escape trial. American Journal of Neuroradiology. 2018;39:102–106

19. Alemseged F, Van Der Hoeven E, Di Giuliano F, Shah D, Sallustio F, Arba F, et al. Response to late-window endovascular revascularization is associated with collateral status in basilar artery occlusion. Stroke. 2019;50:1415–1422

20. Almekhlafi MA, Kunz WG, McTaggart RA, Jayaraman MV, Najm M, Ahn SH, et al. Imaging triage of patients with late-window (6–24 hours) acute ischemic stroke: A comparative study using multiphase ct angiography versus ct perfusion. American Journal of Neuroradiology. 2020;41:129–133

21. Jovin TG, Nogueira RG, Lansberg MG, Demchuk AM, Martins SO, Mocco J, et al. Thrombectomy for anterior circulation stroke beyond 6 h from time last known well (aurora): A systematic review and individual patient data meta-analysis. The Lancet. 2022;399:249–258

22. Desai SM, Aghaebrahim A, Siegler JE, Monteiro A, Jha RM, Jadhav AP, et al. Duration of ischemia impacts postreperfusion clinical outcomes independent of follow-up infarct volume. Stroke: Vascular and Interventional Neurology. 2022;2:e000153

23. Leischner H, Brekenfeld C, Meyer L, Broocks G, Faizy T, McDonough R, et al. Study criteria applied to real life—a multicenter analysis of stroke patients undergoing endovascular treatment in clinical practice. Journal of the American Heart Association. 2021;10

24. Broocks G, Hanning U, Bechstein M, Elsayed S, Faizy TD, Brekenfeld C, et al. Association of thrombectomy with functional outcome for patients with ischemic stroke who presented in the extended time window with extensive signs of infarction. JAMA Network Open. 2022;5:e2235733–e2235733

25. Wollenweber FA, Tiedt S, Alegiani A, Alber B, Bangard C, Berrouschot J, et al. Functional outcome following stroke thrombectomy in clinical practice. Stroke. 2019;50:2500–2506

